# Transdiagnostic MRI Markers of Psychopathology following Traumatic Brain Injury: A Systematic Review and Meta-Analysis Protocol

**DOI:** 10.1101/2023.01.17.23284697

**Authors:** Alexia Samiotis, Amelia J Hicks, Jennie Ponsford, Gershon Spitz

**Author notes:** Registration ID: CRD42022358358 (PROSPERO). 89 Bridge Road Richmond, VIC, 3121. **Author Contributions Alexia Samiotis:** Conceptualization, Methodology, Writing – original draft, Writing – review & editing. **Amelia J Hicks:** Conceptualization, Methodology, Supervision, Writing – review & editing. **Jennie Ponsford:** Conceptualization, Funding acquisition, Methodology, Project administration, Supervision, Writing – review & editing. **Gershon Spitz:** Conceptualization, Methodology, Project administration, Supervision, Writing – review & editing. **Funding statement** This research received no specific grant from any funding agency in the public, commercial or not-for-profit sectors. **Data statement** Statistical code will be available from the Open Science Framework (OSF).

## Abstract

**Introduction:** Psychopathology following traumatic brain injury (TBI) is a common and debilitating consequence that is often associated with reduced functional and psychosocial outcomes. There is a lack of evidence regarding the neural underpinnings of psychopathology following TBI, and whether there may be transdiagnostic neural markers that are shared across traditional psychiatric diagnoses. The aim of this systematic review and meta-analysis is to examine the association of MRI-derived markers of brain structure and function with both transdiagnostic and specific psychopathology following moderate-severe TBI.

**Methods and analysis:** A systematic literature search of Embase (1974–2022), Ovid MEDLINE (1946–2022) and PsycINFO (1806–2022) will be conducted. Publications in English that investigate MRI correlates of psychopathology characterised by formal diagnoses or symptoms of psychopathology in closed moderate-severe TBI populations over 16 years of age will be included. Publications will be excluded that: a) evaluate non-MRI neuroimaging techniques (CT, PET, MEG, EEG); b) comprise primarily a paediatric cohort; c) comprise primarily penetrating TBI. Eligible studies will be assessed against a modified Joanna Briggs Institute Critical Appraisal Instrument and data will be extracted by two independent reviewers. A descriptive analysis of MRI findings will be provided based on qualitative synthesis of data extracted. Quantitative analyses will include a meta-analysis and a network meta-analysis where there is sufficient data available.

**Ethics and dissemination:** Ethics approval is not required for the present study as there will be no original data collected. We intend to disseminate the results through publication to a high-quality peer-reviewed journal and conference presentations on completion.

**PROSPERO registration number:** CRD42022358358

**Article Summary:** Strengths and limitations of this study:

- This is a comprehensive review of MRI markers of psychopathology among adults with moderate – severe traumatic brain injuries.
- We will investigate neural correlates across the spectrum of psychopathology rather than focusing on specific diagnoses, allowing for transdiagnostic investigations of brain structure and function alterations after TBI with comorbid psychopathology.
- We will be restricting eligible studies to English language.
- We will capture pre-injury psychopathology where data are available and analyse the associations with post-injury psychopathology and neural correlates.

## Introduction

Traumatic brain injury (TBI) is a major cause of death, disability, and economic burden worldwide [1]. TBI is defined as an alteration to brain function, or the presence of brain pathology caused by an extraneous force to the head [2]. Closed-head injuries are the most common form of TBI amongst civilians, and occur due to a blunt impact, such as from traffic accidents, falls, and sporting-related injuries [3,4]. These injuries are highly heterogeneous and occur across a spectrum of severity from mild to severe based typically on duration of loss of consciousness, depth of coma and duration of post-traumatic amnesia [5]. Following a TBI, individuals often experience a range of physical, neurological, cognitive, behavioral, functional, and psychological sequelae [6–9]. Psychological sequelae are often the most persistent and disabling in the long-term, particularly for those with moderate – severe injuries [10].

Psychopathology following TBI is both highly prevalent and poorly understood. Approximately 60% of individuals with a moderate – severe injury are diagnosed with a psychiatric disorder within the first year post-injury [11]. Up to 56% of individuals develop a novel psychiatric disorder within the first five years [10]. A significant proportion of individuals have a chronic vulnerability to psychopathology; novel-diagnoses were shown up to 30-years post-injury [12]. The most common psychological conditions post-TBI include mood disorders (43%) and anxiety disorders (21%;[13]), followed by substance use disorders (11.8%; [14]). A significant proportion of individuals also present with a range of symptoms related to emotional distress, which, although disabling, do not meet criteria for a formal diagnosis [11,15]. Despite the prevalence of psychopathology after TBI, psychological conditions are cited as the most unmet need in TBI rehabilitation [16]. Therefore, a better understanding of psychopathology after TBI is required. A wide range of factors have been explored in previous literature to understand psychological outcome, such as premorbid psychiatric history, socioeconomic status, educational attainment, level of early rehabilitation, and psychological adjustment to a life-changing event [10,17–20]. There is also a preliminary but growing body of research investigating whether neural correlates may help to understand the mechanisms involved in psychopathology after TBI [21].

Neuroimaging techniques have been employed to better understand the biological mechanisms underpinning TBI–related psychopathology [21]. Magnetic resonance imaging (MRI) is the most commonly used neuroradiological technique to assess altered brain structure and function. Previous systematic reviews have attempted to integrate MRI findings from studies within TBI populations. These investigations, however, focus on specific disorders. Reviews of depressive disorders [22], anxiety disorders [23], obsessive-compulsive disorders [24] and PTSD [25] showed that regional associations found within individual studies could not be reliably synthesised [22–25].

The lack of consensus on reliable neuroimaging markers of psychopathology after TBI may reflect the significant heterogeneity of injury after TBI, but also could be attributed to limitations of categorical psychopathology. For example, mixed neurobiological findings may arise due to the heterogeneity of symptoms within diagnostic groups [26,27]. This means that two individuals presenting with the same diagnosis could theoretically have no overlapping symptoms. Therefore, different neural substrates could be associated with clusters of symptoms within the same diagnoses, with some preliminary evidence of biologically-driven subgroups in depression [28] and PTSD [29] in non-TBI populations. Another issue with the categorical system is the high comorbidity across diagnostic groups, which suggests that a shared neurobiological substrate may be implicated across diagnostic groups [30,31]. To better understand the neural underpinnings of psychopathology and overcome the shortcomings of categorical classification systems, research in clinical populations has taken a transdiagnostic approach. This approach has been used to assess the mechanisms of a general factor of psychopathology (p-factor). Reviews in adult populations found preliminary evidence for transdiagnostic neural substrates of general psychopathology, and showed that there is difficulty dissociating disorder-specific brain regions or networks [32–36]. For example, reductions in cortical volume converge on the ventromedial prefrontal cortex, medial orbitofrontal cortex, inferior temporal, dorsal, anterior cingulate, and the insula in MDD [37], schizophrenia [38], bipolar disorder [39], and anxiety disorders [32], demonstrating a significant shared diathesis of DSM diagnoses. To date, these transdiagnostic neural substrates have not been examined within a review of psychopathology after TBI.

## Rationale

The neuroscientific literature to date has not identified consistent markers of psychopathology after closed moderate-severe TBI. Inconsistent findings may be due to reliance on categorical classifications of psychopathology and the limited MRI evidence reported to-date.

## Objectives

The objective of the systematic review and meta-analysis is to identify and appraise all studies of MRI markers (structure, microstructure and function) of psychopathology following a closed moderate – severe traumatic brain injury in adults. We aim to synthesise both shared and distinct neural mechanisms to understand whether transdiagnostic MRI markers exist for psychopathology after TBI.

## Methods

This protocol follows the Preferred Reporting Items for Systematic Review and Meta-Analysis Protocols 2015 statement [40] and has been registered at the PROSPERO International Prospective Register of Systematic Reviews of the University of York (CRD42022358358).

### Eligibility criteria

#### Types of studies

All relevant published observational studies (including cohort and case control studies), regardless of sample size, that use MRI to examine brain morphometry (i.e., volume, thickness), white matter microstructure, or functional MRI in at least one area of psychopathology will be included. We will include randomised controlled trials or intervention studies that use MRI to examine brain morphometry, microstructure or function in the context of psychopathology at baseline. The following study types will be excluded from this review: methodological papers, editorials and opinion pieces, qualitative research, individual case reports, and secondary studies such as narrative reviews, systematic reviews and meta-analyses.

#### Types of participants

The eligible population will comprise people aged 16 years and over, of any gender, with a medically confirmed or self-reported diagnosis of moderate or severe closed-head TBI (including complicated mild TBI), as well as any of the following psychiatric conditions: depressive disorders, bipolar disorders, anxiety disorders, post-traumatic stress disorder, acute stress disorder, adjustment disorders, obsessive-compulsive disorders, somatoform disorders, eating disorders, substance use disorders, schizophrenia spectrum and psychotic disorders and personality disorders, or reporting any symptoms of psychopathology.

We will include studies with mixed paediatric and adult populations where the sample consists of at least 80% of people over 16 years or results are presented separately for adults. We will include studies with populations of mixed TBI and other acquired brain injuries where more than 80% of the sample have a TBI or results are presented separately for the TBI group. Severity of TBI will be discerned by medical or self-reports of loss of consciousness of greater than at least 10 minutes or other associated clinical features (i.e., GCS < 13; PTA > 24hrs; positive neuroimaging) as well as structured interview tools (e.g., Ohio State University TBI Identification Method) or questionnaires completed by a clinician or the individual (.

Diagnoses which have been made using standardised diagnostic criteria such as the Diagnostic and Statistical Manual of Mental Disorders, Third Edition (DSM-III), DSM-III-R, DSM-IV, DSM-5 [41], International Classification of Disease, 10^th^ Revision (ICD-10), ICD-11 [42] will be included. We will also include psychopathology captured by self-rating scales such as the Depression Anxiety Stress Scales [43] and the Hospital Anxiety and Depression Scale [44]. Participants with pre-existing psychiatric histories prior to their TBI, as well as those who exhibit psychopathology in more than one area, will be considered eligible. Reports of psychopathology that are made by the participant or clinician will be accepted, however reports by other proxies (i.e., carer, family member) will be excluded.

##### Comparators

The current review will include studies both with and without a control group.

#### Outcome measures

The primary outcomes of the included studies should be structural or functional brain alterations of TBI patients with at least one psychological condition or symptoms in one area of psychopathology. For each study, we plan to obtain information on whole brain and regional-specific measures of the following:

1. Structure: cortical and subcortical gray matter density, thickness, area, or volume.
2. White matter microstructure/integrity: fractional anisotropy, mean diffusivity, axial diffusivity, radial diffusivity.
3. Function: Voxel and region level task-based activation and fMRI based on BOLD signal). Brain network architecture based on resting-state fMRI (rs-fMRI).

Other measures of brain morphometry, white matter microstructure, and function will also be considered.

### Search strategy and data management

#### Search strategy

An electronic systematic search will be conducted across the Embase (1974 – 2022), Ovid MEDLINE (1946 – 2022) and PsycINFO (1806 – 2022) databases in collaboration with an information specialist. The search strategy will include Medical Subject Headings (MeSH) terms, as well as free text, joined with appropriate Boolean operators. The selected search terms are broad, with high sensitivity for identification of relevant studies. All studies in English and published between the above dates and October 2022 will be eligible. A full search strategy for MEDLINE has been provided as an example in Online Supplementary Materials n I.

In addition to the database search, grey literature will be searched using key words in Google Scholar and Research Gate. Experts in the field will be chosen and contacted by the research team.

#### Data management

All citations will be imported into Covidence (https://www.covidence.org) where all review data will be stored and managed.

### Data acquisition and analysis

#### Selection process

After duplicates are removed, the titles and abstracts of articles will be independently reviewed by three reviewers (A.S., G.S. and A.H.) to assess eligibility for inclusion in this review. Eligible citations will be retrieved in full and assessed by two independent reviewers (A.S, & G.S.), with disagreement adjudicated by A.H. and J.P. Where necessary, additional information will be sought from study authors to determine eligibility. Throughout the study selection process, reviewers will not be blinded to the journal titles, study authors or their institutions.

#### Data extraction

Two independent reviewers (A.S. and G.S.) will doubly extract data using a customised data extraction tool based on the standardised tool from the Joanna Briggs Institute System for the Unified Management, Assessment and Review of Information (JBI-SUMARI)[45]. The following information will be retrieved and extracted from each record.

- Basic study identifying information: title, first author, year of publication and country.
- Details of methodology: design (case-control, cohort), participants, sample size, definition/measurement of TBI, TBI severity (mild, moderate or severe), TBI occurrence (single or multiple), demographic characteristics (mean age, time post-injury, handedness, ethnicity and education), population (civilian, military, sport), type of imaging modality (structural MRI/functional MRI), power of the MRI magnetic field (0.35, 0.5, 1.5 or 3 Tesla), metric used (e.g., thickness, fractional anisotropy), rest/active condition (fMRI), brain segmentation process (manual, automatic), psychopathology outcome measure, data analysis strategies.
- Results/Key findings: anatomic locations (x, y, z) based on peak Montreal Neurological Institute (MNI) or Talairach coordinates, cluster size and statistical threshold (Z - statistics, t - statistics, uncorrected p - values) of ROIs analyses that are conducted in the studies; morphological statistics from significant ROIs (e.g., volume, thickness); DTI parameters from significant ROIs; maximum activation of the significant ROIs; the value of clinical characteristics (mean and standard deviation of psychological rating scale scores); and the correlations between imaging data and clinical data.

Any missing information or questions about the above data will be obtained by directly contacting the authors. If no clarification is provided after four weeks, the study will be included in the final analysis with the missing information marked.

### Outcomes and prioritisation

The primary outcomes are MRI markers of psychopathology which may include brain structure, microstructure, and function and these will be prioritised based on technique and psychopathology category. We will prioritise cortical and subcortical structural findings as well as white matter integrity based on DTI. Next, we will focus on cortical and subcortical functional activation patterns based on fMRI. We will categorise data by a) transdiagnostic patterns of brain alterations and b) specific psychopathology-related brain alterations. We will capture pre-injury psychopathology where data are available (e.g., number of studies, study similarities) and analyse the impact on post-injury psychopathology and neural correlates.

### Quality assessment

There are no standard tools for quality appraisal of neuroimaging studies. Therefore, we have devised a customised quality assessment checklist adapted from pre-existing tools: the Joanna Brigg’s Institute Critical Appraisal Tools for cohort studies and for case-control studies, tools used in pre-existing neuroimaging systemic reviews [46,47] and the Committee on Best Practices in Data Analysis Sharing in Neuroimaging MRI (http://www.humanbrainmapping.org). This tool has been included in Online Supplementary Materials Appendix II for reference. Two reviewers (A.S. and G.S.) will independently score the included articles against the checklist and study quality will be defined as high (> 8 criteria met), medium (5 – 8) or low (< 5) for Category 1 and high (> 5 criteria met), medium (3 – 5) or low (< 3) for Category 2.

### Data synthesis

Data extracted from studies will be summarised in a table, including basic study identifying information, details of methodology and results. A qualitative review of the structural and functional brain correlates will be performed and synthesised into a detailed narrative of findings. First, we will present a narrative synthesis of neural correlates which are identified as common to multiple domains of psychopathology, structured by neuroimaging modality (structural MRI, DTI, fMRI). Next, findings will be integrated by domain of psychopathology (e.g., depressive disorders, anxiety disorders), structured by neuroimaging modality.

### Meta-analysis

If there are a sufficient number of studies available (i.e., at least two) that analyse a domain of psychopathology using a similar methodology, then we will use meta-analysis. Where feasible, we will first obtain separate effect sizes for each study. Next, we will estimate Hedge’s g for whole brain and region of interest measurements including volumetric, morphometric, white matter integrity and activation results. We will utilise a random effects model to pool the effect sizes. Meta-regression analyses will consider the following potential covariates: sample size, mean age, % of females, methodological quality scores, year of publication, years-post injury and GCS score/duration of PTA. Where data is sparse, we will use a beta-binomial model for meta-analysis (see Mathes and Kuss, 2017). We will use R studio, R packages “meta” and “metasens” following the guide outlined in Balduzzi et al. (2019).

Heterogeneity will be assessed using prediction intervals (generated from the T statistic) where there is a sufficient number of studies or minimal heterogeneity across pooled studies, and from the I-squared statistic [48].

### Network meta-analysis

We will conduct a network meta-analysis where there is sufficient data available and sufficient similarity across study characteristics. We will employ the “netmeta” package in R for a frequentist approach to synthesising biological data across studies [49]. The aim of our network meta-analysis will be to understand commonalities between brain structure and function across different psychopathology in TBI studies. We will visualise associations across studies using a network plot, where each node is reflective of a particular psychopathology (e.g., depressive disorders and associated symptomatology), the node size relative to the number of studies included and the edges between nodes relative to the number of pairwise comparisons. Hedge’s g will be utilised to understand the brain structure and function differences between psychopathology classifications.

### Meta-bias(es)

We will assess for the possibility of small-study effects and publication bias in our meta-analysis by inspecting funnel plot asymmetry and Egger’s test (p < 0.1); following Balduzzi et al. (2019)[50].

### Confidence in cumulative evidence

The quality of evidence for primary outcomes will be assessed using the Grading of Recommendations Assessment, Development and Evaluation (GRADE) guidelines [51].

### Patient and Public Involvement

There are currently no plans to involve patients or the public in the design, conduct, or reporting, or dissemination of our systematic review.

## Conclusion

No clear and reliable neuroimaging markers of psychopathology have been identified to date, possibly due to the significant reliance on categorical classification systems. The present review utilises a transdiagnostic approach to investigate the possibility that shared neural substrates converge across the TBI literature more reliably than psychopathology-specific substrates. This paper summarises the protocol for a systematic review and meta-analysis which will synthesise both distinct and shared neural mechanisms of psychopathology across neuroimaging studies in moderate-severe TBI. This in turn will provide a greater understanding of biological mechanisms which may contribute to the expression of psychopathology after a TBI.

## Data Availability

Statistical code will be available from the Open Science Framework (OSF).

## Acknowledgements

Our information specialist, Farhad Shokraneh for his contribution in design of the search strategies and collaboration with the search.

