## Supplementary material for "Transdiagnostic MRI Markers of Psychopathology following Traumatic Brain Injury: A Systematic Review and Meta-Analysis Protocol": Online Supplementary Materials

**Title**

**Appendix I**

**Search strategy for MEDLINE**

***B. MEDLINE***

Database: Ovid MEDLINE(R) ALL <1946 to October, 2022>

1	Brain Concussion/ OR Brain Contusion/ OR Brain Damage, Chronic/ OR Brain Injuries, Traumatic/ OR Brain Injuries/ OR Brain Injury, Chronic/ OR Brain Stem Hemorrhage, Traumatic/ OR Cerebral Hemorrhage, Traumatic/ OR Cerebral Hemorrhage/ OR Cerebrovascular Trauma/ OR Diffuse Axonal Injury/ OR exp Brain Hemorrhage, Traumatic/ OR exp Brain Injuries, Diffuse/ OR exp Hematoma, Subdural/ OR exp Intracranial Hemorrhages/ OR Head Injuries, Closed/ OR Head Injuries, Penetrating/ OR Hematoma, Epidural, Cranial/ OR Hematoma, Subdural, Acute/ OR Hematoma, Subdural, Chronic/ OR Hematoma, Subdural, Intracranial/ OR Intracranial Hemorrhage, Traumatic/ OR Skull Fractures/ OR Subarachnoid Hemorrhage, Traumatic/ OR Subarachnoid Hemorrhage/ OR Trauma, Nervous System/ OR (Traumatic OR Posttrauma* OR Brain Arachnoid Haemorrhage* OR Brain Arachnoid Hemorrhage* OR Brain Bleeding* OR Brain Bruise* OR Brain Contus* OR Brain Damage* OR Brain Haemorrhage* OR Brain Hemorrhage* OR Brain Injur* OR Brain Lesion* OR Brain System Trauma* OR Brain Trauma* OR Brain Vascular Injur* OR Brain Vascular Trauma* OR Broken Skull* OR Cerebral Bleeding OR Cerebral Concuss* OR Cerebral Contus* OR Cerebral Damage* OR Cerebral Haematoma* OR Cerebral Haemorrhage* OR Cerebral Hematoma* OR Cerebral Hemorrhage* OR Cerebral Injur* OR Cerebral Lesion* OR Cerebral Trauma* OR Cerebrocranial Injur* OR Cerebrocranial Trauma* OR Cerebrovascular Trauma* OR Cerebrum Haemorrhage* OR Cerebrum Hemorrhage* OR Cerebrum Lesion* OR Commotio* OR Concuss* OR DAI OR DAIs OR Damaged Brain* OR Depressed Cranium* OR Depressed Skull* OR Diffuse Axonal Injur* OR Forebrain Lesion* OR Fracture Skull* OR Fractured Cranial Bone* OR Fractured Cranium* OR Fractured Skull* OR Frontal Region Trauma* OR Haemisphere Injur* OR Haemisphere Lesion* OR Head Bruis* OR Head Contus* OR Head Injur* OR Head Trauma* OR Head Wound* OR Hemisphere Injur* OR Hemisphere Lesion* OR Interhemispheric Haematoma* OR Interhemispheric Hematoma* OR Intracerebral Bleeding* OR Intracerebral Haematoma* OR Intracerebral Haemorrhage* OR Intracerebral Hematoma* OR Intracerebral Hemorrhage* OR Intracortical Haemorrhage* OR Intracortical Hemorrhage* OR Intracranial Bleeding* OR Intracranial Epidural Haematoma* OR Intracranial Epidural Hematoma* OR Intracranial Haematoma* OR Intracranial Haemorrhage* OR Intracranial Hematoma* OR Intracranial Hemorrhage* OR Intracranial Lesion* OR Intraventricular Haemorrhage* OR Intraventricular Hemorrhage* OR Midbrain Haemorrhage* OR Midbrain Hemorrhage* OR mTBI OR mTBIs OR Nervous System Injur* OR Nervous System Trauma* OR Neuroinjur* OR Neuro-Injur* OR Neurologic Damage* OR Neurotrauma* OR Occipital Region Trauma* OR Occipital Trauma* OR Parietal Region Trauma* OR Skull Depressed Fracture* OR Skull Fracture* OR Skull Frontobasal Fracture* Skullbase Fracture* OR Subarachnoid Bleeding* OR Subarachnoid Haematoma* OR Subarachnoid Haemorrhage* OR Subarachnoid Haemorrhagia* OR Subarachnoid Hematoma* OR Subarachnoid Hemorrhage* OR Subarachnoid Hemorrhagia* OR Subarachnoidal Bleeding* OR Subarachnoidal Haemorrhage* OR Subarachnoidal Hemorrhage* OR Subdural Bleeding* OR Subdural Haematoma* OR Subdural Haemorrhage* OR Subdural Hematoma* OR Subdural Hemorrhage* OR Subepidural Haematoma* OR Subepidural Hematoma* OR Temporal Region Trauma* OR TBI OR TBIs).ti,ab.

2	Psychopathology/ OR Mental Disorders/ OR exp Mood Disorders/ OR Cyclothymic Disorder/ OR exp Depressive Disorder/ OR Depressive Disorder, Major/ OR Depressive Disorder, Treatment-Resistant/ OR Dysthymic Disorder/ OR Premenstrual Dysphoric Disorder/ OR Seasonal Affective Disorder/ OR Mental Fatigue/ OR exp Anxiety Disorders/ OR Anxiety, Separation/ OR Neurocirculatory Asthenia/ OR Neurotic Disorders/ OR exp Obsessive-Compulsive Disorder/ OR Hoarding Disorder/ OR Panic Disorder/ OR exp Phobic Disorders/ OR Phobia, Social/ OR Stress Disorders, Post-Traumatic/ OR Adjustment Disorders/ OR exp Somatoform Disorders/ OR Body Dysmorphic Disorders/ OR Body Integrity Identity Disorder/ OR Conversion Disorder/ OR Factitious Disorders/ OR Munchausen Syndrome/ OR "Munchausen Syndrome by Proxy"/ OR Hypochondriasis/ OR Neurasthenia/ OR exp "Feeding and Eating Disorders"/ OR Anorexia Nervosa/ OR Avoidant Restrictive Food Intake Disorder/ OR Binge-Eating Disorder/ OR Bulimia Nervosa/ OR Diabulimia/ OR Food Addiction/ OR Night Eating Syndrome/ OR Orthorexia Nervosa/ OR Pica/ OR Rumination Syndrome/ OR exp Substance-Related Disorders/ OR exp Alcohol Amnestic Disorder/ OR Alcoholic Intoxication/ OR Alcoholic Korsakoff Syndrome/ OR Alcoholic Neuropathy/ OR Alcohol-Induced Disorders/ OR Alcohol-Induced Disorders, Nervous System/ OR Alcoholism/ OR Alcohol-Related Disorders/ OR Amphetamine-Related Disorders/ OR Binge Drinking/ OR Cocaine-Related Disorders/ OR Drug Overdose/ OR Heroin Dependence/ OR Inhalant Abuse/ OR Marijuana Abuse/ OR "Marijuana Use"/ OR Morphine Dependence/ OR Narcotic-Related Disorders/ OR Opiate Overdose/ OR exp Opioid-Related Disorders/ OR Opium Dependence/ OR Phencyclidine Abuse/ OR Psychoses, Alcoholic/ OR Psychoses, Substance-Induced/ OR Substance Abuse, Intravenous/ OR Substance Abuse, Oral/ OR "Tobacco Use Disorder"/ OR "Schizophrenia Spectrum and Other Psychotic Disorders"/ OR Schizotypal Personality Disorder/ OR exp Schizophrenia/ OR Schizophrenia, Catatonic/ OR Schizophrenia, Disorganized/ OR Schizophrenia, Paranoid/ OR Schizophrenia, Treatment-Resistant/ OR Shared Paranoid Disorder/ OR Affective Disorders, Psychotic/ OR Capgras Syndrome/ OR Delusional Parasitosis/ OR Paranoid Disorders/ OR Psychotic Disorders/ OR Psychoses, Substance-Induced/ OR Psychoses, Alcoholic/ OR exp Personality Disorders/ OR Antisocial Personality Disorder/ OR Borderline Personality Disorder/ OR Compulsive Personality Disorder/ OR Dependent Personality Disorder/ OR exp Histrionic Personality Disorder/ OR Hysteria/ OR Paranoid Personality Disorder/ OR Passive-Aggressive Personality Disorder/ OR Schizoid Personality Disorder/ OR Schizotypal Personality Disorder/ OR Dissociative Identity Disorder/ OR (Abnormal Mental State* OR Behavior Disorder* OR Diseased Mental State* OR Disordered Mental State* OR Disturbed Mental State* OR Insanity OR Mental Abnormalit* OR Mental Change* OR Mental Confusion* OR Mental Defect* OR Mental Disorder* OR Mental Disturbance* OR Mental Illness* OR Mental Insufficiency OR Mental Symptom* OR Mentally Ill OR Neuropsychiatric Disease* OR Neuropsychiatric Disorder* OR Psychiatric Diagnosis OR Psychiatric Disease* OR Psychiatric Disorder* OR Psychiatric Illness* OR Psychiatric Symptom* OR Psychic Disease* OR Psychic Disorder* OR Psychic Disturbance* OR Psychologic Disorder* OR Psychologic Disturbance* OR Psychological Disorder* OR Psychological Disturbance* OR Psychopatholog* OR Psycho-Patholog* OR Mood Disorder* OR Mood Disturbance* OR Affective Disorder* OR Affective Disturbance* OR Affective Illness* OR Cyclothymi* OR Depress* OR Melancholia* OR Paraphrenia* OR Dysthymi* OR Premenstrual Dysphoric Disorder* OR Premenstrual Dysphoric Syndrome* OR Psychosis OR Psychoses OR Psychotic OR Blunted Affect OR Flat Affect OR Schizo* OR Mania* OR Manic OR Bipolar OR Mental Fatigue OR Hypomani* OR Anxi* OR Astheni* OR Neurotic OR Neurosis OR Neuroses OR Mourning Syndrome OR Obsess* OR Compuls* OR OCD OR Hoarding OR Panic OR Phobic OR Phobia* OR Effort Syndrome OR Psychoneuros* OR Anankastic Personalit* OR Stress* OR Catastrophiz* OR Catastrophis* OR Distress* OR Koro* OR Psychasthenia* OR Repetitive Behaviour* OR Repetitive Behavior* OR Trichotillomania* OR Hair Pulling OR PTSD OR Posttraumatic Psychic Syndrome OR Posttraumatic Syndrome OR Moral Injur* OR Posttraumatic Stress* OR Post-Traumatic Stress* OR Traumatic Stress OR Shell Shock OR Complex Trauma OR DESNOS OR "Disorders of Extreme Stress Not Otherwise Specified" OR Adjustment Disorder* OR Adjustment Reaction* OR Anniversary Reaction* OR Reactive Depression* OR Reactive Disorder* OR Transient Situational Disorder* OR Transient Situational Disturbance* OR Amputee Identity Disorder* OR Apotemnophilia* OR Astasia-Abasia OR BIID OR Bodily Distress Disorder* OR Bodily Distress Disorders OR Body Dysmorphic Disorder* OR Body Image Disfunction* OR Body Image Disorder* OR Body Integrity Disorder* OR Body Integrity Identity Disorder* OR Briquet Syndrome* OR Cardiophobia* OR Conversion Disorder* OR Conversion Hysteri* OR Conversion Reaction* OR Conversion Syndrome* OR Da Costa Syndrome* OR Dysmorphophobia OR Factitious Disorder* OR Foreign Limb Syndrome* OR Functional Heart Complaint* OR Functional Movement Disorder* OR Functional Neurologic Symptom Disorder* OR Functional Neurological Disorder* OR Functional Neurological Symptom Disorder* OR Ganser Syndrome OR Globus OR Hospital Addiction Syndrome* OR Hypochondria* OR Hysteria OR Hysteric* OR Medically Unexplained Syndrome* OR Munchausen Syndrome OR Munchhausen Syndrome OR Neurasthenia OR Neurocirculation Asthenia OR Neurocirculatory Asthenia OR Neurocirculatory Dystonia OR Neurodermatitis OR Neurogenic Heart OR "Pain Disorder" OR Pseudocyesis OR Pseudopsychosis OR Psychalg* OR Psychogenic Amaurosis OR Psychogenic Amblyopia OR Psychogenic Blindness OR Psychogenic Nonepileptic Seizure* OR Psychogenic Non-Epileptic Seizure* OR Psychogenic Pain OR Psychogenic Seizure* OR Psychogenic Vision OR Psychogenic Visual OR Psychophysiologic Disorder* OR Psychosomatic Blindness OR Psychosomatic Disorder* OR Soldier Heart OR Somatic Symptom Disorder* OR Somatization* OR Somatisation* OR Somatoform Disorder* OR Somatoform Pain* OR Xenomelia* OR Feeding Disorder* OR Feeding Disorder OR Eating Disorder* OR Appetite Disorder* OR Disordered Eating OR Eating Pathology OR Pathologic Eating OR Pathologic Feeding OR Pathological Eating OR Pathological Feeding OR Anorexia Nervosa* OR Avoidant Restrictive Food Intake Disorder OR Binge-Eating Disorder OR Bulimia* OR Diabulimia* OR Food Addiction* OR Compulsive Eating OR Night Eating Syndrome* OR Orthorexia OR Pica OR Rumination Syndrome* OR Merycism OR Rumination Disorder* OR ARFID OR Emotional Eating OR Food Aversion OR Food Refusal OR Muscle Dysmorphia OR Purging Disorder* OR Trichophag* OR Hyperphag* OR Polyphag* OR Addict* OR Alcohol Abuse* OR Alcohol Amnes* OR Alcohol Depend* OR Alcohol Induced Amnes* OR Alcohol Induced Disorder* OR Alcohol Induced Dysmnesi* OR Alcohol Induced Korsakoff Syndrome* OR Alcohol Induced Persisting Amnestic Disorder* OR Alcohol Problem* OR Alcohol Related Disorder* OR Alcohol Related Seizure* OR Alcohol Syndrome* OR "Alcohol Use Disorder*" OR Alcoholic* OR Alcoholism OR Amphetamine Abuse* OR Amphetamine Dependen* OR Amphetamine Related Disorder* OR "Amphetamine Use Disorder*" OR Angel Dust Abuse* OR Benzodiazepine Dependen* OR Binge Alcohol* OR Binge Drink* OR Cannabis Abuse* OR Cannabis Dependen* OR Cannabis Related Disorder* OR Chemical Dependen* OR Cocaine Abuse* OR Cocaine Dependen* OR Cocaine Related Disorder* OR Delirium Tremens OR Dipsomani* OR Drug Abuse* OR Drug Bombing OR Drug Craving OR Drug Dependen* OR Drug Facilitation* OR Drug Habituation* OR Drug Misuse* OR Drug Overdose* OR Drug Parachuting OR Drug Physical Dependen* OR Drug Psychoses OR Drug Related Craving OR "Drug Use Disorder*" OR Drugs Craving OR Drunkenness* OR Ethanol Abuse* OR Ethanol Dependen* OR Hashish Abuse* OR Heroin Abuse* OR Heroin Dependen* OR Heroin Smoking* OR Heroinism OR Inhalant Abuse* OR Marihuana Abuse* OR Marijuana Abuse* OR Marijuana Dependen* OR Marijuana Related Disorder* OR "Marijuana Use*" OR Medication Misuse* OR Medicine Misuse* OR Methamphetamine Dependen* OR Morphine Abuse* OR Morphine Dependen* OR Narcotic Abuse* OR Narcotic Dependen* OR Narcotic Related Disorder* OR Narcotism OR Nicotine Abuse* OR Nicotine Dependen* OR "Nicotine Use Disorder*" OR Nicotinism OR Opiate Abuse* OR Opiate Alkaloid Dependen* OR Opiate Crisis OR Opiate Dependen* OR Opiate Epidemic* OR Opiate Overdose* OR Opioid Abuse* OR Opioid Crisis OR Opioid Dependen* OR Opioid Epidemic* OR Opioid Misuse* OR Opioid Overdose* OR Opioid Related Disorder* OR "Opioid Use Disorder*" OR Opioids Crisis OR Opioids Dependen* OR Opioids Epidemic* OR Opium Abuse* OR Opium Dependen* OR Opium Epidemic* OR Opium Smoking OR "Opium Use*" OR PCP Abuse* OR PCP Dependen* OR Phencyclidine Abuse* OR Phencyclidine Dependen* OR Phencyclidine Related Disorder* OR Physical Dependen* OR "Polydrug Use*" OR Substance Abuse* OR Substance Craving OR Substance Dependen* OR Substance Induced Psychoses OR Substance Related Disorder* OR "Substance Use*" OR Tobacco Abuse* OR Tobacco Dependen* OR "Tobacco Use Disorder*" OR Tobaccoism OR Toxic Psychoses OR Toxicomani* OR Paranoi* OR Delus* OR Psychogenic Parasitos* OR Capgras Syndrome* OR Hebephreni* OR Hallucinosis OR Hallucinat* OR Anankastic Personalit* OR Antisocial Behavior* OR Antisocial Behaviour* OR Antisocial Personalit* OR Anti-Social Personalit* OR "As If Personality" OR "As If Personalities" OR Asthenic Personalit* OR Avoidant Personalit* OR Borderline OR Character Deviation* OR Character Disorder* OR Character Disturbance* OR Character Problem* OR Dependent Personalit* OR Diogenes Syndrome OR Dissociative Identity Disorder* OR Dual Personalit* OR Dyssocial Behavior* OR Dyssocial Behaviour* OR Histrionic Personalit* OR Impulse-Ridden Personalit* OR Inadequate Personalit* OR Masochis* OR Multiple Identity Disorder* OR Multiple Personalit* OR Narcism OR Narcissism OR Narcissistic OR Neuropsychopath* OR Passive-Aggressive Personalit* OR Personality Change OR Personality Disorder* OR Personality Disturbance* OR Psychological Dependen* OR Psychopath* OR Sadis* OR Sadomasochis* OR "Self Love" OR Senile Squalor Syndrome OR Social Behavior Disorder* OR Social Breakdown Syndrome OR Social Disease* OR Social Patholog* OR Sociopath*).ti,ab.

3	exp Magnetic Resonance Imaging/ OR exp Neuroimaging/ OR exp Brain Mapping/ OR Neuroradiography/ OR Diffusion Magnetic Resonance Imaging/ OR Diffusion Tensor Imaging/ OR Brain Cortical Thickness/ OR Cerebral Cortical Thinning/ OR White Matter/ OR Gray Matter/ OR Anisotropy/ OR (Magnetic Resonance OR MRI OR MRIs OR MR Imag* OR NMR OR MR Tomogra* OR Zeugmatogra* OR Chemical Shift Imag* OR Magneti?ation Transfer OR Proton Spin Tomogra* OR fMRI OR "Spin Echo" OR Diffusion Imag* OR "Diffusion Tensor" OR DTI OR Tractogra* OR "Diffusion Weighted" OR "Echo Planar" OR Echoplanar OR Neuroimag* OR (Neural adj Imag*) OR Neuro-Imag* OR (Brain adj Imag*) OR Brain Mapping OR Stereotaxic Atlas OR Functional Cerebral Locali?ation? OR Connectom* OR Connectivity Map OR Connectivity Matrix OR Cortical Thickness OR Cortex Thickness OR Grey Matter Thickness OR Neuroradio* OR DWI OR Fluid-Attenuated Inversion Recovery OR FLAIR OR GRE OR Gradient Recalled Echo* OR Gradient Echo* OR Cortical Thinning OR Subcortical Thickness OR Anisotrop* OR Anisotroph* OR Diffusivit* OR Diffusion Rate* OR Gray Matter* OR Grey Matter* OR Voxel Based Morphometr* OR VBM).ti,ab.

4	and/1-3

5	exp Animals/ not Humans.sh.

6	4 not 5

7	limit 6 to undetermined

8	limit 6 to english language

9	7 OR 8

**Appendix II**

**Customised Quality Assessment Tool**

Category 1: sample characteristics

|  | Yes | No | Unclear | Not applicable |
| --- | --- | --- | --- | --- |
| 1. Were the groups comparable other than the presence of disease in cases or the absence of disease in controls? | □ | □ | □ | □ |
| 2. Were cases and controls matched appropriately? | □ | □ | □ | □ |
| 3. Were the same criteria used for identification of cases and controls? | □ | □ | □ | □ |
| 4. Was exposure measured in a standard, valid and reliable way? | □ | □ | □ | □ |
| 5. Important demographic data (age and gender) were reported with mean (or median) and standard deviations (or range)? | □ | □ | □ | □ |
| 6. Were confounding factors identified? | □ | □ | □ | □ |
| 7. Were strategies to deal with confounding factors stated? | □ | □ | □ | □ |
| 8. Were outcomes assessed in a standard, valid and reliable way for cases and controls? | □ | □ | □ | □ |
| 9. Important clinical variables (eg, depressive episodes, depression-related scale score) were reported with mean (or median) and standard deviations (or range)? | □ | □ | □ | □ |
| 10. Sample size per group >10? | □ | □ | □ | □ |

Category 2: methodology and reporting characteristics

|  | Yes | No | Unclear | Not applicable |
| --- | --- | --- | --- | --- |
| 1. Magnet strength at least 1.5T? | □ | □ | □ | □ |
| 2. At least 5 min of resting state acquisition? | □ | □ | □ | □ |
| 3. Whole brain coverage of resting scans? | □ | □ | □ | □ |
| 4. The acquisition and preprocessing techniques were clearly described so that they could be reproduced? | □ | □ | □ | □ |
| 5. Coordinates reported in a standard space? | □ | □ | □ | □ |
| 6. Significant results are reported after correction for multiple comparisons using a standard statistical procedure (e.g., FDR (False Discovery Rate), FWE (Family Wise Error) or permutation-based methods)? | □ | □ | □ | □ |
| 7. Conclusions were consistent with the results obtained and the limitations were discussed? | □ | □ | □ | □ |
